# Paediatric COVID-19 severity across SARS-CoV-2 variants in hospitalised children in South Africa: a retrospective cohort study

**DOI:** 10.64898/2026.07.06.26357377

**Authors:** Stefania Fiandrino, Costanza Di Chiara, Daniele Donà, Rory Dunbar, Pierre Goussard, Harsha Lochan, Helena Rabie, Andrew Redfern, Carien Truter, Margaret Van Niekerk, Gert van Zyl, Lilly M. Verhagen, Marieke van der Zalm, Daniela Paolotti

## Abstract

The evolving epidemiology of COVID-19, driven by successive SARS-CoV-2 variants of concern (VOCs), has prompted ongoing evaluation of their impact on disease severity in children. In low-and middle-income countries (LMICs), children experience a higher burden of severe respiratory illness and pneumonia-related mortality due to factors such as malnutrition, incomplete immunisation, HIV exposure or infection, tuberculosis, and disparities in access to healthcare services. Hospital-based paediatric studies from LMICs are therefore needed to understand how the epidemiology and severity of COVID-19 have changed across pandemic waves. This study examined 354 hospitalised children with SARS-CoV-2 infection during the ancestral, pre-Omicron (Beta and Delta), and Omicron waves at Tygerberg Hospital in Cape Town, South Africa. We analysed data collected over an extended period, from March 2020 to June 2022. Statistical analyses were used to describe clinical characteristics across variant periods, and multivariable logistic regression models were applied to evaluate associations between potential risk factors and disease severity. Paediatric COVID-19 severity varied across VOC periods, with the highest burden observed during the pre-Omicron (Beta and Delta) waves. In multivariable analyses, younger age and circulating variants were associated with disease severity; CRP levels emerged as a marker associated with more severe illness, and corticosteroid treatment, while also associated with disease severity, reflects clinical response to more severe cases. These findings contribute to a better understanding of the epidemiology and clinical impact of COVID-19 in children and highlight the importance of context-specific surveillance and treatment strategies in resource-limited settings.

**What is already known on this topic:** The epidemiology and clinical characteristics of COVID-19 evolved substantially with successive SARS-CoV-2 variants of concern, yet their impact on disease severity in children remains unclear. Existing evidence is largely limited to adult populations, children with mild-to-moderate illness, or well-resourced healthcare settings, leaving a critical gap in knowledge on how epidemiology and disease severity changed across pandemic waves among hospitalised children in low- and middle-income countries.

**What this study adds:** While hospital-based studies in South Africa have substantially advanced understanding of paediatric SARS-CoV-2 infection, they have largely been limited to specific pandemic periods, individual variants, or specific levels of care. By analysing data over an extended period (March 2020 to June 2022), this study provides a systematic characterisation of changes in clinical presentation and risk factors for severe disease across the ancestral, pre-Omicron (Beta and Delta), and Omicron waves in hospitalised children.

**How this study might affect research, practice or policy:** These findings underscore the need for age-stratified, variant-aware surveillance systems in low- and middle-income countries, where the burden of severe paediatric respiratory illness remains high. Access to oxygen in these settings also needs to be addressed. The association between young age and severe outcomes supports the prioritisation of infants in clinical monitoring and triage protocols. The prognostic value of CRP as an early risk stratification biomarker and the use of corticosteroids highlight the need for paediatric-specific trials to establish evidence-based treatment protocols in resource-limited settings.

## 1 Introduction

Since the emergence of severe acute respiratory syndrome coronavirus 2 (SARS-CoV-2), children have generally experienced a milder clinical course of COVID-19 than adults, with lower susceptibility to infection and reduced risks of symptomatic disease, hospitalisation, intensive care unit (ICU) admission, and mortality [1–3]. Nevertheless, severe disease occurs in a subset of paediatric patients, particularly young children and infants, some of whom require intensive care, respiratory support, or prolonged hospitalisation [4, 5]. Evidence on determinants of severe COVID-19 in paediatric patients remains limited and heterogeneous, particularly in hospitalised populations. Hospital-based studies identify associations with prolonged symptoms, elevated inflammatory markers, hypoxemia at presentation, and need for mechanical ventilation [6–9]. Reported risk factors vary across studies, with some identifying older age as a predictor of critical illness, while others report greater severity among younger children, including infants. Multicountry analyses further demonstrate age-dependent differences in disease severity over the course of the pandemic [10]. These changes reflect the evolving epidemiology of COVID-19, driven by successive SARS-CoV-2 variants of concern (VOCs) with differing virulence and transmissibility. However, the impact of VOCs on disease severity and clinical manifestations in children remains unclear. To date, most studies on the clinical impact of SARS-CoV-2 variants have focused on adult populations, children with mild to moderate disease, or well-resourced healthcare settings [11–14]. In contrast, children in low- and middle-income countries (LMICs) experience a higher burden of severe respiratory disease and pneumonia-related mortality, driven by factors including malnutrition, incomplete immunisation, HIV exposure or infection, tuberculosis, air pollution, poverty, and disparities in access to healthcare services [15, 16]. Consequently, paediatric studies from LMICs are needed to characterise changes in epidemiology and disease severity across pandemic waves. Hospital-based studies from Cape Town, in Africa, have substantially advanced understanding of paediatric SARS-CoV-2 infection, describing clinical presentation and outcomes across pandemic phases and levels of care [17–20]. Collectively, these studies show that infants are overrepresented among paediatric hospital admissions and frequently present with severe respiratory illness requiring advanced support, particularly early in the pandemic [17]. Subsequent analyses from Tygerberg Hospital and district-level facilities report greater morbidity and longer oxygen requirements in SARS-CoV-2–associated illness compared with other acute respiratory infections, with young age, HIV exposure, prematurity, and tuberculosis co-infection common among hospitalised children [18, 20]. During the Omicron wave, infants and children with comorbidities remained most affected, with no clear increase in severity among children living with HIV or tuberculosis [19]. Despite providing important insights, these studies were largely limited to specific pandemic periods, individual variants, or specific levels of care. Consequently, changes in clinical presentation and risk factors of disease severity across successive SARS-CoV-2 variants of concern have not been systematically evaluated within a single hospital-based paediatric population over time. To address this gap, this study examines changes in clinical characteristics and identifies risk factors for severe disease among children hospitalised with SARS-CoV-2 infection across the ancestral, pre-Omicron (Beta and Delta), and Omicron waves at Tygerberg Hospital, a major secondary and tertiary referral center in Cape Town, South Africa. We use statistical analyses to characterise clinical features of COVID-19 across different VOCs and multivariable logistic regression models to assess associations between risk factors and disease severity.

## 2 Materials and Methods

### 2.1 Settings and study population

This study presents routine care data from the COVID-Kids cohort in South Africa, including hospi­talised children aged 0–15 years with a laboratory-confirmed diagnosis of SARS-CoV-2 who presented to Tygerberg Hospital (TBH) between March 18, 2020, and June 17, 2022. TBH is a secondary and tertiary referral hospital in Cape Town, serving a population with significant socio-economic disadvan­tages and a high burden of infectious diseases, including tuberculosis (TB) and HIV. Infants diagnosed in the neonatal service and children with multisystem inflammatory syndrome (MIS-C) who did not have a positive SARS-CoV-2 rRT-PCR result were excluded. The final study cohort comprised 354 children. The study was approved by the Human Research Ethics Committee of the Faculty of Health Sciences, Stellenbosch University, South Africa (HREC N20/04/013_COVID). The analysis used rou­tinely collected data that were entered without patient identifiers. A waiver of informed consent was granted.

### 2.2 Patient and public involvement

Patients and members of the public were not involved in the design, conduct, reporting, or dissemi­nation plans of this research. This study analysed routinely collected clinical data from hospitalised children as part of the COVID-Kids cohort in South Africa. Given the retrospective nature of the study and the use of anonymised data, direct patient involvement in the development of research questions, outcome measures, or dissemination strategy was neither feasible nor appropriate.

### 2.3 Study Definitions

#### 2.3.1 Variables

For each child admitted with SARS-CoV-2, clinicians providing routine care prospectively completed a standardised case report form. Clinical assessments were conducted on admission, including the collection of demographic characteristics (age, sex, date of birth), anthropometric measurements (weight and height), clinical presentation (reported symptoms, vital signs, and physical examination), medical assessment (hydration status), past medical history and comorbidities (including tuberculosis, HIV infection, asthma, chronic lung disease, congenital cardiac disorders, autoimmune disorders, diabetes, and malignancy), and details of hospital management. Treatment-related data included both COVID-19–specific treatments (e.g., corticosteroids) and non–COVID-19–related treatments (e.g., oral or intravenous antibiotics), as well as the need for supplemental oxygen and admission to the paediatric ICU. Laboratory investigations, such as C-reactive protein (CRP) and other relevant tests, were also recorded. SARS-CoV-2 variant classification and disease severity were assigned for each patient. SARS-CoV-2 variant labels were assigned, distinguishing between Ancestral, Beta, Delta, and Omicron waves. For this analysis, Beta and Delta variants were combined into a single pre-Omicron category. This approach reflects the sequential dominance of Beta and Delta variants before the emergence of Omicron, as well as shared epidemiological characteristics, including associations with higher viral loads and shifts toward younger age groups among infected individuals in South Africa [21]. Also, Delta infection has been associated with relatively greater disease severity in paediatric populations compared with Omicron, further supporting the use of a consolidated pre-Omicron category for comparative analyses across distinct phases of the pandemic [22]. Accordingly, we delineated the groups as follows: Ancestral (March 18 – October 4, 2020), Pre-Omicron (November 1, 2020 – October 7, 2021), which considers the period during which the Beta and Delta variants co-circulated, and Omicron (November 17, 2021 – June 17, 2022).

For analysis, children were categorised into three age groups: *<* 1 year, 1–4 years, and *≥* 5 years. Nutritional status was classified as underweight or obese based on weight- and height-for-age using the Centers for Disease Control and Prevention (CDC) growth references and was included among comorbidities [23]. Tuberculosis was classified as either current or previous disease. HIV infection was defined as confirmed HIV-positive status; children with a CD4 count *<* 200 *cells/µL* additionally stratified to account for the severity of immunosuppression. Respiratory support was categorized as non-invasive ventilation (including nasal prongs, high-flow nasal cannula (HFNC), and continuous positive airway pressure (CPAP)) or invasive mechanical ventilation.

Clinician-assigned disease severity was classified according to the adapted World Health Organization (WHO) severity grading system for SARS-COV-2 infection [19]: mild illness included symptomatic children without lower respiratory involvement or oxygen requirement; moderate illness involved lower respiratory disease without hypoxia; severe illness was defined by hypoxia or significant respiratory distress; and critical illness by respiratory failure, shock, or multi-organ dysfunction. This information was included only in stratified statistical analyses to explore differences in clinician-assigned disease severity across SARS-CoV-2 variants.

#### 2.3.2 Outcomes

The primary outcome was severe COVID-19 outcome, defined as a composite outcome of paediatric ICU admission, invasive mechanical ventilation, or in-hospital death, consistent with definitions used in international guidelines and in previous paediatric COVID-19 studies [24–26]. A secondary outcome was paediatric ICU admission, analysed separately, as the number of events for invasive mechanical ventilation and in-hospital death was insufficient to support separate adjusted models. We note that this outcome definition is distinct from the WHO-based disease severity classification assigned by clinicians.

### 2.4 Statistical Analysis

Baseline demographic, clinical characteristics, and COVID-19–related outcomes were summarised using frequencies and proportions for categorical variables, and medians with interquartile ranges (IQRs) for continuous variables. Comparisons between variant of concern (VOC) groups were performed using the ² test for categorical variables, or the Fisher–Freeman–Halton exact test when any expected cell frequency was below 5; continuous variables were compared using the Kruskal–Wallis test.

Associations between potential risk factors and COVID-19 severity were assessed using multivari­able logistic regression models. Two binary outcomes were examined: admission to the paediatric ICU and severe COVID-19 outcome. Variables were selected a priori based on clinical relevance, and adjusted models controlled for demographic factors (age and sex), SARS-CoV-2 variant (Ancestral, pre-Omicron, Omicron), and clinical markers reflecting disease status, including severe dehydration, corticosteroid treatment, and CRP levels as a marker of inflammation. Adjusted odds ratios (aORs) with 95% confidence intervals (CIs) were reported. Missing data were handled using complete-case analysis. A p-value *<* 0.05 was considered statistically significant. Statistical analyses were conducted using Python (version 3.9.6).

## 3 Results

Among the 354 children included in the analysis, the median age was 1.2 years (IQR: 0.3–5.3), and 170 (48%) were female. Sixteen children (5%) were living with HIV, 41 (12%) had a diagnosis of tuber­culosis, and 101 (29%) presented with at least one co-existing medical condition. Clinicians classified disease severity as mild in 45%, moderate in 30%, severe in 9%, and critical in 7% of cases. Admissions varied substantially over time, peaking in August 2021 and again in December 2021–January 2022, with the highest proportions of moderate, severe, and critical cases coinciding with these surge periods (Figure 1).

**Figure 1:**
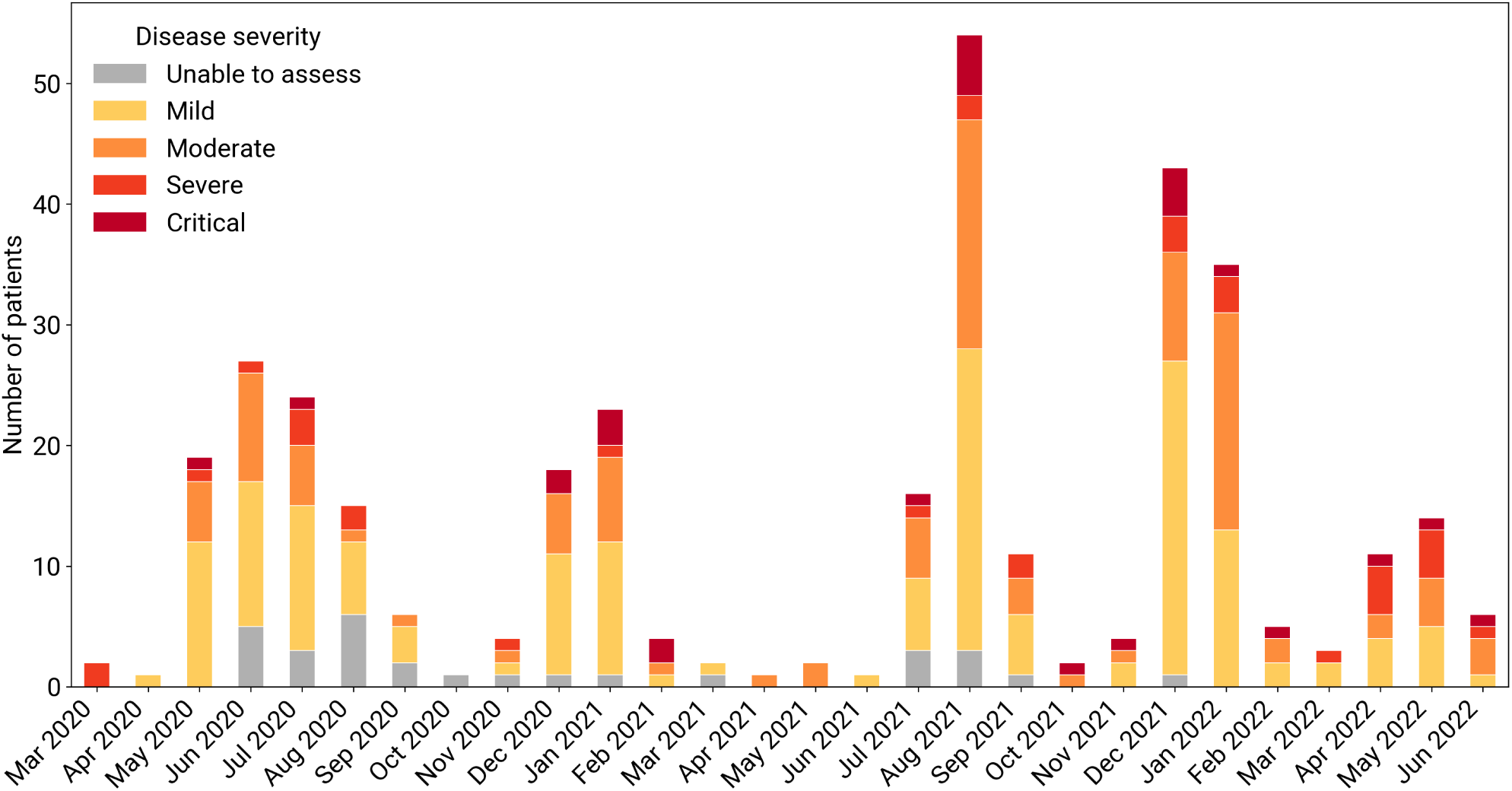
Monthly COVID-19 hospital admissions stratified by clinician-assigned disease severity (March 2020 - June 2022). Each bar represents the total number of admissions in a given month, with colours indicating the proportion of each severity category.

Descriptive statistics are reported by variant of concern (VoC) period. Three periods have been considered, based on the VoC labels provided in the dataset: the Ancestral period includes 95 hospi­talised patients, the pre-Omicron period, including Beta and Delta variants, includes 138 hospitalised patients, and the Omicron period includes 121 hospitalised patients. Table 1 and Table 2 summarise clinical symptoms, laboratory investigations, and treatments by variant group. Table 3 details clinical outcomes across the variant-defined periods.

**Table 1:** Sociodemographic characteristics of COVID-19 hospitalised cases by variant of concern. Dif­ferences between VoC groups were analyzed using the *χ*^2^ test or Fisher-Freeman-Halton exact test for categorical variables (with Fisher-Freeman-Halton applied when expected cell frequencies were <5), and the Kruskal-Wallis test for continuous variables.

| Characteristic | Total<br>n = 354 | Ancestral<br>n = 95 | Pre-Omicron<br>n = 138 | Omicron<br>n = 121 | p-value |
| --- | --- | --- | --- | --- | --- |
| **Age (yr), median (IQR)** | 1.2 (0.3–5.3) | 1.2 (0.2–4.0) | 1.6 (0.4–5.9) | 1.0 (0.3–6.9) | 0.31 |
| **Age, n (%)** |  |  |  |  |  |
| < 1 yr | 158 (44.6) | 43 (45.3) | 57 (41.3) | 58 (47.9) | 0.34 |
| 1–4 yr | 87 (24.6) | 28 (29.5) | 36 (26.1) | 23 (19.0) |  |
| > 5 yr | 109 (30.8) | 24 (25.3) | 45 (32.6) | 40 (33.1) |  |
| **Sex, n (%)** |  |  |  |  |  |
| Female | 170 (48.0) | 46 (48.4) | 71 (51.4) | 53 (43.8) | 0.47 |
| Male | 184 (52.0) | 49 (51.6) | 67 (48.6) | 68 (56.2) |  |
| **Comorbidities, n (%)** |  |  |  |  |  |
| Obesity | 3 (0.9) | 1 (1.1) | 0 (0.0) | 2 (1.7) | 0.37 |
| Underweight | 8 (2.3) | 1 (1.1) | 2 (1.4) | 5 (4.1) | 0.32 |
| Asthma | 6 (1.7) | 2 (2.1) | 3 (2.2) | 1 (0.8) | 0.94 |
| Chronic lung disease | 6 (1.7) | 2 (2.1) | 2 (1.4) | 2 (1.7) | 0.99 |
| Autoimmune disorder | 8 (2.3) | 2 (2.1) | 6 (4.3) | 0 (0.0) | 0.20 |
| Cardiac disorder | 11 (3.1) | 3 (3.2) | 3 (2.2) | 5 (4.1) | 0.86 |
| Diabetes | 3 (0.9) | 0 (0.0) | 1 (0.7) | 2 (1.7) | 0.84 |
| Cerebral palsy | 8 (2.3) | 0 (0.0) | 5 (3.6) | 3 (2.5) | 0.43 |
| Malignancy | 17 (4.8) | 7 (7.4) | 5 (3.6) | 5 (4.1) | 0.62 |
| Others | 64 (18.1) | 17 (17.9) | 26 (18.8) | 21 (17.4) | 0.95 |
| TB infected | 41 (11.6) | 8 (8.4) | 16 (11.6) | 17 (14.0) | 0.44 |
| HIV infected | 16 (4.5) | 2 (2.1) | 5 (3.6) | 9 (7.4) | 0.19 |
| Any comorbidity | 101 (28.5) | 26 (27.4) | 38 (27.5) | 37 (30.6) | 0.83 |

**TB current infection:** 6 (6.3%) in Ancestral, 15 (10.9%) in pre-Omicron, and 12 (9.9%) in Omicron period; $p = 0.48$ . **TB previous infection:** 2 (2.1%) in Ancestral, 1 (0.7%) in pre-Omicron period, and 5 (4.1%) in Omicron period; $p = 0.18$ .
**CD4 < 200 cells/ μL:** 0 (0.0%) in Ancestral, 1 (0.7%) in Pre-Omicron, and 3 (2.5%) in Omicron period; $p = 0.24$ .

**Table 2:**
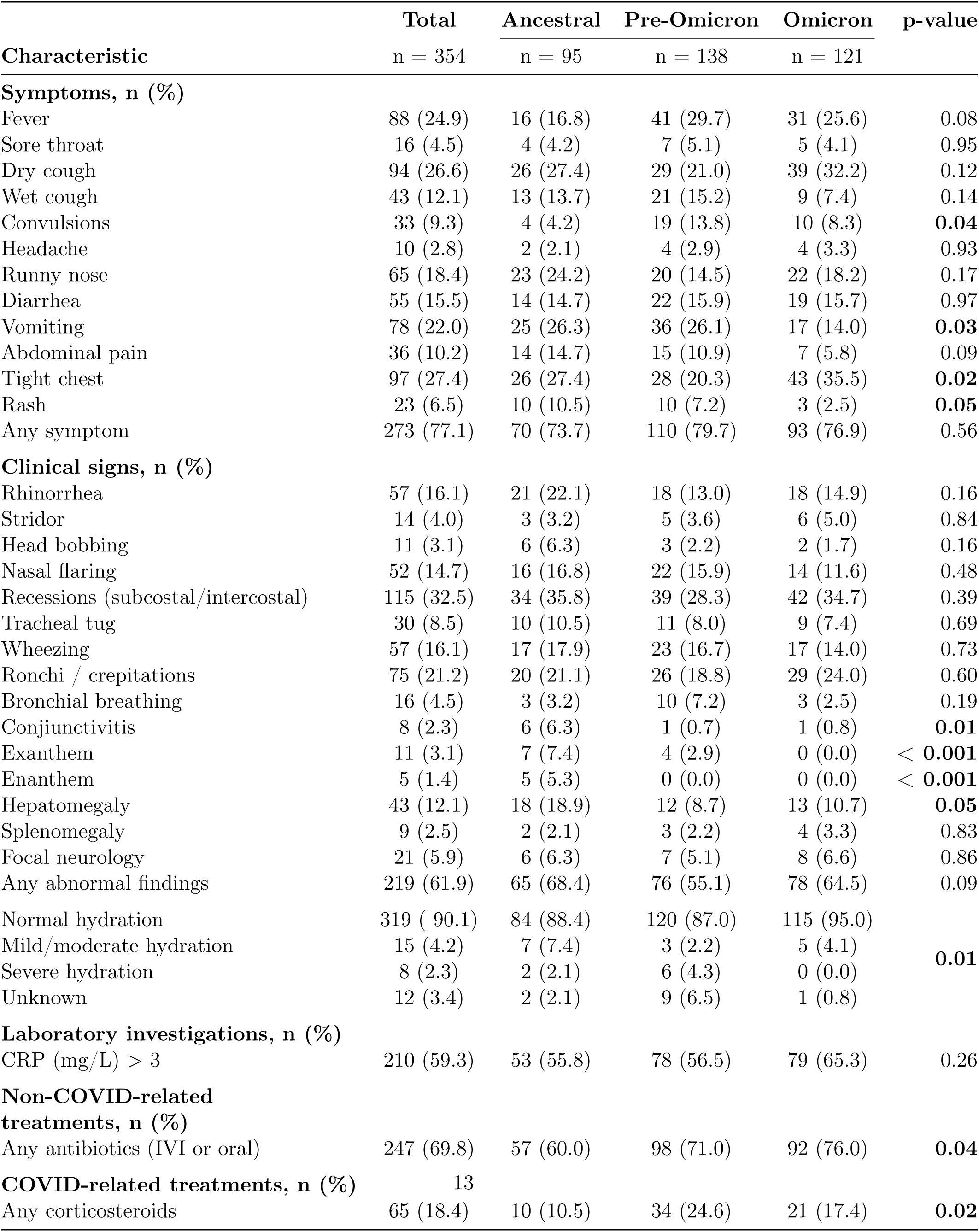
Clinical symptoms and treatments of COVID-19 hospitalised cases by variant of concern. Differences between VoC groups were analyzed using the *χ*^2^ test or Fisher-Freeman-Halton exact test for categorical variables (with Fisher-Freeman-Halton applied when expected cell frequencies were <5), and the Kruskal-Wallis test for continuous variables.

**Table 3:** Outcomes of COVID-19 hospitalised cases by variant of concern. Differences between VoC groups were analyzed using the *χ*^2^ test or Fisher-Freeman-Halton exact test for categorical variables (with Fisher-Freeman-Halton applied when expected cell frequencies were <5), and the Kruskal-Wallis test for continuous variables.

| Characteristic | Total<br>n = 354 | Ancestral<br>n = 95 | Pre-Omicron<br>n = 138 | Omicron<br>n = 121 | p-value |
| --- | --- | --- | --- | --- | --- |
| **Respiratory support, n (%)** |  |  |  |  |  |
| **Non-invasive ventilation** |  |  |  |  |  |
| Nasal prongs | 73 (20.6) | 11 (11.6) | 35 (25.4) | 27 (22.3) | **0.01** |
| High flow | 13 (3.7) | 4 (4.2) | 4 (2.9) | 5 (4.1) |  |
| CPAP | 19 (5.4) | 5 (5.3) | 3 (2.2) | 11 (9.1) |  |
| **Invasive ventilation** |  |  |  |  |  |
| Ventilated | 25 (7.1) | 2 (2.1) | 13 (9.4) | 10 (8.3) |  |
| **Severity of disease, n (%)** |  |  |  |  |  |
| Mild | 162 (45.8) | 46 (48.4) | 61 (44.2) | 55 (45.5) | < **0.001** |
| Moderate | 105 (29.7) | 21 (22.1) | 45 (32.6) | 39 (32.2) |  |
| Severe | 32 (9.0) | 9 (9.5) | 7 (5.1) | 16 (13.2) |  |
| Critical | 26 (7.3) | 2 (2.1) | 14 (10.1) | 10 (8.3) |  |
| Unable to assess | 29 (8.2) | 17 (17.9) | 11 (8.0) | 1 (0.8) |  |
| **Outcomes, n (%)** |  |  |  |  |  |
| Invasive ventilation | 25 (7.1) | 2 (2.1) | 13 (9.4) | 10 (8.3) | 0.08 |
| Paediatric ICU | 71 (20.1) | 14 (14.7) | 37 (26.8) | 20 (16.5) | **0.04** |
| Child died | 12 (3.4) | 2 (2.1) | 7 (5.1) | 3 (2.5) | 0.43 |
| Severe COVID-19 outcome | 78 (22.0) | 15 (15.8) | 42 (30.4) | 21 (17.4) | **0.01** |
| **Length of hospital stay (d), median (IQR)** | 6.0 (2.0–12.0) | 5.0 (2.0–14.0) | 6.0 (2.0–13.0) | 6.0 (3.0–10.0) | 0.84 |

Overall, we observed no statistically significant differences in sociodemographic characteristics across the Ancestral, pre-Omicron, and Omicron periods(Table 1). Several clinical features showed significant variation by variant (Table 2). These include symptoms such as convulsions, tight chest, rash, and vomiting, as well as physical signs like conjunctivitis, exanthem, enanthem, and hepatomegaly. Notably, hydration status also differs significantly between groups. In the pre-Omicron cohort, 87% of children had normal hydration, whereas 4.3% exhibited severe dehydration. Conversely, in the Omicron cohort, 95% maintained normal hydration, and none experienced severe dehydration. Both non-COVID-related and COVID-related treatment types vary across the variant-defined periods, and differences are statistically significant (Table 2). COVID-related treatment (corticosteroids) was most common in the pre-Omicron period (approximately 25%), followed by a lower rate during the Omicron period (17%) and the lowest during the Ancestral period (about 10%). Non–COVID-related treatment (antibiotics) showed a consistent increasing trend from the Ancestral period through to Omicron, with the highest frequency observed during the Omicron period.

Several outcomes of COVID-19 differed significantly by VOCs. Among hospitalised children, the need for respiratory support varied significantly by variant period (p= 0.01, Table 3), although the difference in invasive ventilation alone did not reach significance (p = 0.08): in the Ancestral period approximately 21% of children required non-invasive respiratory support (including nasal prongs, high-flow, or CPAP), while about 2% needed invasive ventilation; in the pre-Omicron period the proportion needing non-invasive support increased to nearly 30%, and over 9% required invasive ventilation, and in the Omicron period the non-invasive support was administered to nearly 35% of children, with invasive ventilation necessary for around 8%. The highest proportion of critical cases occurred during the pre-Omicron period (10% of cases during the period), followed by the Omicron period (8%), while only 2% of cases during the Ancestral period presented with critical disease. Similarly, paediatric ICUs were highest in the pre-Omicron cohort (approximately 27%), followed by the Omicron period (around 16%), and the Ancestral period (14%). Overall mortality reached more than 3%, with the highest death rate observed during the pre-Omicron phase, although the difference across variant periods was not statistically significant (p = 0.43). The percentage of children meeting this composite severity outcome was highest in the pre-Omicron period (30%), compared to approximately 17% during Omicron and about 16% in the Ancestral period.

To address our second research objective, we constructed two multivariable logistic regression models to identify factors associated with COVID-19 severity outcomes. We first examined a baseline model including demographic and epidemiological risk factors (age, gender, SARS-CoV-2 variant). Then, we test a second clinical model, additionally incorporating clinical markers measured during hospitalisation (CRP levels, severe dehydration status) and the corticosteroid use, to assess their independent prognostic value. The baseline model (Figure 2A-B) included demographic and epidemiological factors available at presentation (age group, gender, SARS-CoV-2 variant). For paediatric ICU (Figure 2A), children aged 1-4 years old had lower odds compared to infants aged 0-1 years [*aOR* = 0.44, 95% *CI* : 0.22 *−* 0.91*, p <* 0.05], while infection with a pre-Omicron variant was associated with increased odds compared to the Ancestral period [*aOR* = 2.16, 95%*CI* : 1.08 *−* 4.30*, p <* 0.05]. For the composite severe COVID-19 outcome (Figure 2B), only pre-Omicron infection remained significantly associated with severity [*aOR* = 2.36, 95% *CI* : 1.21 *−* 4.59*, p <* 0.05].

**Figure 2:**
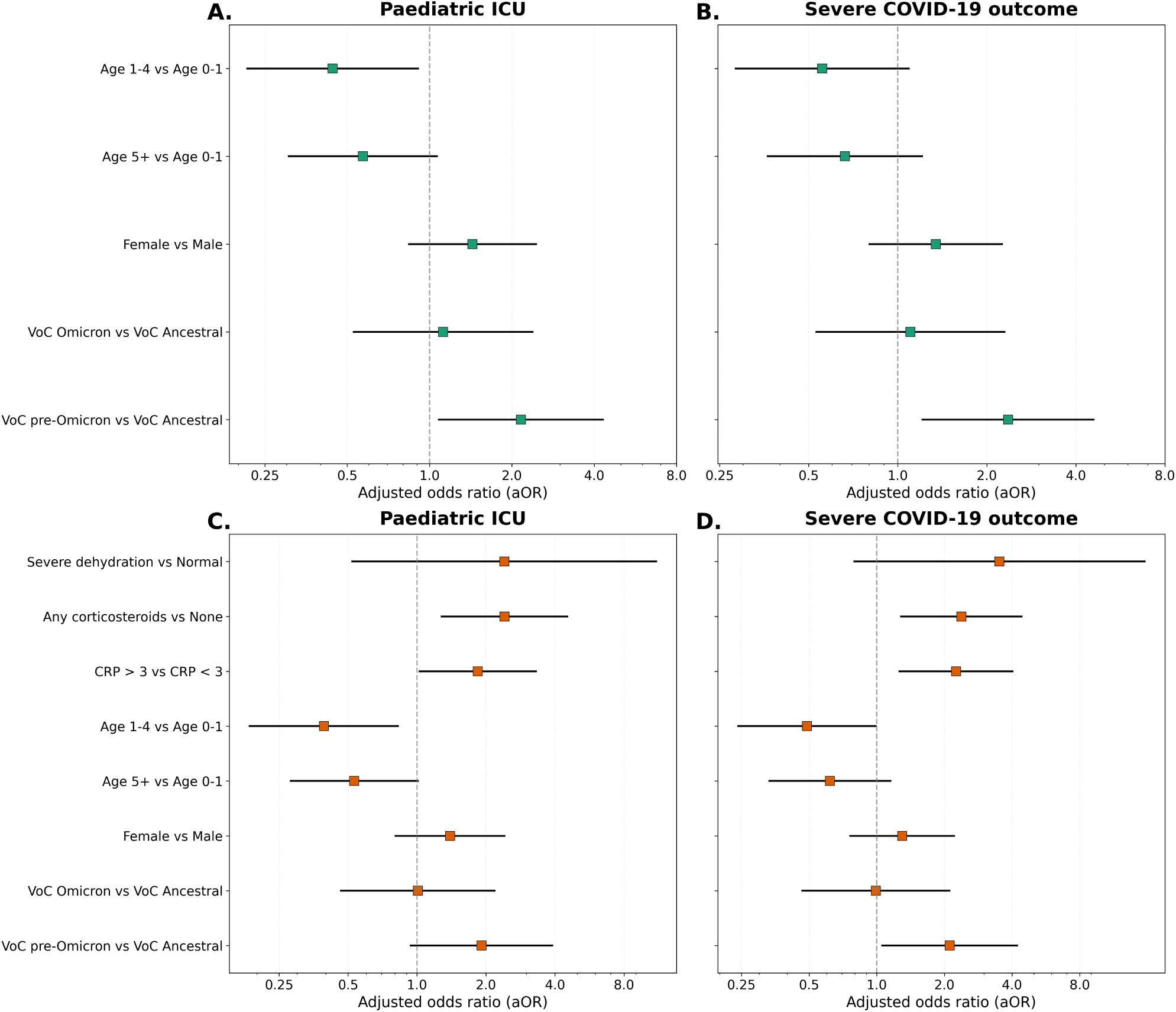
Multivariable logistic regression models for paediatric ICU admission and severe COVID-19. Forest plots showing adjusted odds ratios (aOR) with 95% confidence intervals for factors associated with paediatric ICU (left column) and severe COVID-19 outcome (right column). (A-B) Baseline models including demographic and epidemiological risk factors only (age group, gender, SARS-CoV-2 variant). (C-D) Full models additionally incorporate clinical markers measured during hospitalisation (C-reactive protein, corticosteroid use, severe dehydration status). Green squares and error bars represent point estimates and 95% CIs for baseline models; orange squares error bars represent point estimates and 95% CIs for full models. The vertical dashed line indicates an odds ratio of 1 (no association). Values are plotted on a logarithmic scale.

The second model included also clinical markers as CRP level and severe dehydration status, and the corticosteroid use, given its clinical relevance and differential administration across disease severity levels (Figure 2C-D). For paediatric ICU admission (Figure 2C), children aged 1-4 years continued to show lower odds compared to infants [*aOR* = 0.39, 95% *CI* : 0.19*−*0.83*, p <* 0.05], while corticosteroid treatment was strongly associated with increased odds [*aOR* = 2.41, 95% *CI* : 1.28 *−* 4.53*, p <* 0.01]. For severe COVID-19 (Figure 2D), elevated CRP levels were associated with increased odds of severe COVID-19 [*aOR* = 2.23, 95% *CI* : 1.21 *−* 4.13*, p* = 0.01]; corticosteroid treatment was similarly associated with a greater likelihood of severe disease [*aOR* = 2.38, 95% *CI* : 1.28 *−* 4.41*, p <* 0.01]; infection with a pre-Omicron variant was also linked to higher odds of severe COVID-19 compared to patients infected in the Ancestral period [*aOR* = 2.11, 95% *CI* : 1.05 *−* 4.22*, p <* 0.05], while children aged 1-4 years old had lower odds compared to infants aged 0-1 years [*aOR* = 0.49, 95% *CI* : 0.24 *−* 0.99*, p <* 0.05].

## 4 Discussion

In this study, we characterise changes in clinical characteristics of COVID-19 across different VOCs, and we evaluate the risk of severe COVID-19 outcomes among hospitalised children in South Africa. Our findings contribute to the growing literature on COVID-19 in children, with a focus on middle-income countries. We found that infants aged 0-1 years had higher odds of both paediatric ICU and severe COVID-19 outcome, aligning with the existing literature [16, 27, 28]. Growing evidence indicates that young age is an important risk factor for severe SARS-CoV-2 infection. Bhuiyan et al. [27] found that half of COVID-19 cases in very young children occurred in infants. Similarly, Graff et al. [28] reported that infants under 3 months of age were more likely to be hospitalised with COVID-19. In addition, a large study conducted across six sub-Saharan African countries [16], which also included children from the present analysis, showed young age as a key predictor of severe disease.

A previous study conducted in a South African cohort reported that infections with the Beta and Delta variants were more common among younger individuals compared with the ancestral variant, across age groups ranging from early childhood to older adulthood [21]. Our findings are consistent with this evidence, showing that individuals infected during the pre-Omicron period were more likely to experience severe COVID-19 than those infected with the ancestral variant. In interpreting the higher severity observed during the pre-Omicron period, several mechanisms should be considered. First, accumulating evidence suggests that the Beta and Delta variants were intrinsically more virulent than earlier lineages. Infections with these variants have been associated with higher peak viral loads, potentially increasing infectiousness [21], as well as with increased risks of severe outcomes, including higher incidence of COVID-19, more rapid increases in hospital admissions, and higher in-hospital mortality [29]. Together, these findings support a biological explanation in which variant-specific pathogenicity contributed to the observed differences in disease severity. However, the observed variant-period effect is unlikely to be attributable to viral biology alone. The pre-Omicron waves in South Africa occurred in a context of lower population immunity, characterised by limited prior infection and no vaccination among children, which likely increased susceptibility to severe disease. Several studies\ consistently reported markedly lower clinical severity during the Omicron-dominated wave compared with preceding variants [30–32]. These studies suggest that the reduced severity observed among hospitalised patients during the Omicron period was likely driven by a combination of a less virulent virus and high levels of population immunity resulting from previous infections [30]. Accordingly, the lower severity during the Omicron wave probably reflected decreased intrinsic virulence, increased reinfections occurring in the presence of pre-existing immunity, although the relative contributions of these factors remain difficult to disentangle [33]. Finally, analyses from South Africa [34] have shown that health system pressure varied substantially over time, influencing access to intensive care, triage practices, and the composition of hospitalised patients. During periods of high demand, stricter admission and triage criteria resulted in a more selective hospitalised case mix, whereas eased capacity in later periods was associated with the admission of milder cases. These temporal changes in health-system context may have contributed to differences in observed disease severity across pandemic waves, underscoring that such differences are unlikely to reflect viral characteristics alone and remain difficult to disentangle from biological and immunological factors.

C-reactive protein (CRP) is produced by the liver in response to infection, tissue damage, or inflammation, and elevated CRP levels reflect the acute-phase response. In our analysis, higher CRP levels were observed in children presenting with severe COVID-19, consistent with prior studies in adults and children [35, 36]. This association likely reflects greater systemic inflammation in more severe disease rather than CRP being a causal risk factor. This finding reinforces the value of CRP as a prognostic biomarker to aid clinical assessment and risk stratification in pediatric COVID-19.

Evidence from adult populations suggests that corticosteroid treatment can reduce mortality in critically ill COVID-19 patients [37]. A large meta-analysis of clinical trials showed a beneficial effect of systemic corticosteroids in adults; however, these studies were conducted exclusively in high-income settings and did not include children. In our analysis, corticosteroid use was positively associated with severe COVID-19, likely reflecting confounding by indication: corticosteroids were preferentially given to the sickest children. This does not suggest harmful treatment effects, but rather that corticosteroid use marks disease severity. This does not contradict the meta-analysis findings, as the association we observed is driven by disease severity at the time of treatment, not by the treatment effect. Nevertheless, this highlights the need for further studies to better understand the role and optimal use of corticosteroids in paediatric COVID-19 and subsequent viral pandemics, particularly in resource-limited settings. Future work could explore inference methods such as propensity score matching or weighting to better account for systematic differences in baseline characteristics, reduce the effects of confounding, and more accurately estimate the effect of corticosteroids on clinical outcomes.

This study comes with several limitations that should be considered when interpreting the findings. First, the analysis was based on routine care data from a single tertiary referral hospital, which may limit the generalizability of the results to other settings, particularly primary care or non-hospitalised paediatric populations. As Tygerberg Hospital provides secondary and tertiary care, the study population likely represents children with more severe illness, and the overall burden and spectrum of COVID-19 severity in the community may therefore be underestimated. This selection bias, however, was not uniform across the study period: during the early phase, the Tygerberg Hospital was a designated COVID-19 facility centralising cases across the full severity spectrum, meaning that less severe presentations were also captured. Second, the observational design precludes causal inference. Associations between risk factors and severe COVID-19 may be influenced by residual confounding, despite adjustment for key demographic factors, clinical variables, and markers of disease severity. Third, the sample size and number of severe outcomes were limited, preventing separate adjusted models for rare outcomes such as invasive mechanical ventilation and death. Finally, some potentially relevant variables, such as vaccination status, socioeconomic indicators at the individual level, detailed immunological markers, seasonal variation, and information regarding co-infection, were not consistently available in routine clinical records and could not be included in the analysis.

In conclusion, this study analysed data covering a period from March 2020 to June 2022, with the strength of capturing shifts in the clinical characteristics of COVID-19 across different variants in a middle-income country context. We evaluated the association between COVID-19 risk factors and disease severity outcomes, confirming the role of age and circulating variants in shaping disease severity, as well as the prognostic role of CRP as a marker of disease severity. These findings improve\ understanding of COVID-19 epidemiology in children and underscore the need to tailor monitoring and treatment strategies to local, resource-limited settings. Beyond its immediate clinical implications, this study provides important insights for future pandemic preparedness. Our findings highlight the value of prospective, variant-informed surveillance systems in paediatric hospital settings. Timely collection and analysis of data on how pathogen evolution influences disease severity and clinical outcomes can support the adaptation of clinical guidelines and public health strategies. Looking ahead, the early implementation of standardized data collection protocols will enable robust longitudinal and variant-stratified analyses, ultimately supporting more responsive, evidence-driven clinical care and policy decision-making in future health emergencies.

## 5 Declarations

### 5.2 Authors’ contributions

Stefania Fiandrino: Writing – review & editing, Writing – original draft, Software, Methodology, Investigation, Formal analysis. Costanza Di Chiara: Writing – review & editing, Methodology. Daniele Donà: Writing – review & editing, Methodology. Rory Dunbar: Writing – review & editing, Data curation. Pierre Goussard: Writing – review & editing, Data curation. Harsha Lochan: Writing – review & editing, Data curation. Helena Rabie: Writing – review & editing, Data curation. Andrew Redfern: Writing – review & editing, Data curation. Carien Truter: Writing – review & editing, Data curation. Margaret Van Niekerk: Writing – review & editing, Data curation. Gert van Zyl: Writing – review & editing, Data curation. Lilly M. Verhagen: Writing – review & editing, Data curation. Marieke van der Zalm: Writing – review & editing, Data curation, Investigation, Supervision, Conceptualization. Daniela Paolotti: Writing – review & editing, Data curation, Investigation, Supervision, Conceptu­alization. All authors approved the final manuscript as submitted, and accepted responsibility for submitting it for publication.

## Data Availability

All data produced in the present study are available upon reasonable request to the authors.

